# Effectiveness of Ivermectin as add-on Therapy in COVID-19 Management (Pilot Trial)

**DOI:** 10.1101/2020.07.07.20145979

**Authors:** Faiq I Gorial, Sabeeh Mashhadani, Hend M Sayaly, Basim Dhawi Dakhil, Marwan M. AlMashhadani, Adnan M Aljabory, Hassan M Abbas, Mohammed Ghanim, Jawad I Rasheed

## Abstract

**Background:** To date no effective therapy has been demonstrated for COVID-19. In vitro, studies indicated that ivermectin (IVM) has antiviral effect.

**Objectives:** To assess the effectiveness of ivermectin (IVM) as add-on therapy to hydroxychloroquine (HCQ) and azithromycin (AZT) in treatment of COVID-19.

**Methods:** This Pilot clinical trial conducted on hospitalized adult patients with mild to moderate COVID-19 diagnosed according to WHO interim guidance. Sixteen Patients received a single dose of IVM 200Mcg /kg on admission day as add on therapy to hydroxychloroquine (HCQ)and Azithromycin (AZT) and were compared with 71 controls received HCQ and AZT matched in age, gender, clinical features, and comorbidities.

The primary outcome was percentage of cured patients, defined as symptoms free to be discharged from the hospital and 2 consecutive negative PCR test from nasopharyngeal swabs at least 24 hours apart. The secondary outcomes were time to cure in both groups and evaluated by measuring time from admission of the patient to the hospital till discharge.

**Results:** Of 87 patients included in the study,t he mean age ± SD (range) of patients in the IVM group was similar to controls [44.87 ± 10.64 (28-60) vs 45.23 ± 18.47 (8-80) years, p=0.78] Majority of patients in both groups were male but statistically not significant [11(69%) versus 52 (73%), with male: female ratio 2.21 versus 2.7-, p=0.72)

All the patients of IVM group were cured compared with the controls [16 (100 %) vs 69 (97.2 %)]. Two patients died in the controls. The mean time to stay in the hospital was significantly lower in IVM group compared with the controls (7.62 ± 2.75 versus 13.22 ±5.90 days, p=0.00005, effect size= 0.82). No adverse events were observed

**Conclusions:** Add-on use of IVM to HCQ and AZT had better effectiveness, shorter hospital stay, and relatively safe compared with controls. however, a larger prospective study with longer follow up may be needed to validate these results.

## Introduction

A novel coronavirus, severe acute respiratory syndrome coronavirus 2 (SARS-CoV-2), was first identified in December 2019 as the cause of a respiratory illness designated coronavirus disease 2019, or Covid-19 with significant public health impact (1). Several therapeutic agents have been evaluated for the treatment of Covid-19, however, none have yet been shown to be effective (2,3)

Recently some reports on HCQ [4-6], Azithromycin [7] and Ivermectin [8] have shown therapeutic effects against novel coronavirus infection. Ivermectin is an antiparasitic drug with a broad spectrum antiviral effect Recently, in vitro study showed reduction of viral RNA in Vero-hSLAM cells 2 hours postinfection with SARS-CoV-2 clinical isolate Australia/VIC01/2020 (8). The authors hypothesized that the effect was likely due to the inhibition of IMP α/β1-mediated nuclear import of viral proteins.

Because of the broad spectral antiviral activities of IVM and it is safety profile, It may offer a therapeutic potential to COVID-19. This study was designed to assess effectiveness and safety of add-on use of IVM to HCQ and AZT in COVID 19 patients.

## Patients and Methods

### Study design

This pilot interventional single center study with a synthetic controlled arm (SCA) was conducted at Al-Shifa’a Hospital Center from first of April to the end of May 2020. Synthetic controlled arm was used due to difficulty of using placebo for our patients and the strong preference for the investigational product in this pandemic Covid-19 disease to improve drug development and reduce patients burden. SCA is an external control constructed from patient-level data from previous patients records to match the baseline characteristics of the patients in an investigational group and augment a single-arm trial to estimate treatment effects. The SCA in this trial included previous patients who were treated by HCQ and AZT according to the Iraqi Ministry of Health protocols for treatment of covid-19.

Ethical approval of the study was taken in accordance with the Declaration of Helsinki and its amendments and the Guidelines for Good Clinical Practices issued by the Committee of Propriety Medicinal Product of the European Union from Iraqi ministry of health and the study was registered with No. 497 at April 2020. Also, this study was registered in ClinicalTrials.gov website under identifier number: NCT04343092. Informed consent was obtained from the participants to admit the study.

## Participants

### Inclusion criteria

Inclusion criteria were the following: 1) men and women with age at least 18 years 2) mild to moderate COVID-19 diagnosed by positive polymerase chain reaction (PCR) testing <=3 days from enrollment 3)Patient acceptance and willingness to comply with planned study procedures and to complete the follow up. 4) hospital admission 5) no participation in other clinical trials, such as antiviral trials, during the study period. 6) Able to provide informed consent

Mild and moderate COVID-19 were defined according to World Health Organization (WHO) interim guidance (16). Mild COVID-19 was defined as symptomatic patients meeting the case definition for COVID-19 without evidence of viral pneumonia or hypoxia. The symptoms included: fever, cough, fatigue, anorexia, shortness of breath, myalgias. Other non specific symptoms such as soar throat, nasal congestion, headache, diarrhea nausea, vomiting, loss of smell, loss of taste, Older people and immunosuppressed patients in particular may present with atypical symptoms such as fatigue, reduced alertness, reduced mobility, diarrhea, loss of appetite, delirium, and absence of fever. Moderate COVID-19: included adolescent or adult with clinical signs of pneumonia (fever, cough, dyspnea, fast breathing) but no signs of severe pneumonia, including SpO2 ≥ 90% on room air.

### Exclusion criteria

Exclusion criteria were the following: 1) severe COVID-19 defined as respiratory distress (≥ 30 breaths/min; in resting state, oxygen saturation of 93% or less on room air; or arterial partial pressure of oxygen (PaO2)/fraction of inspired oxygen (FIO2) of 300 or less. 2) Life threatening COVID-19 was defined as respiratory failure requiring mechanical ventilation; shock; or other organ failure (apart from lung) requiring intensive care unit (ICU) monitoring. 3) hypersensitivity or severe adverse events to IVM, 4) Alanine Aminotransferase (ALT) or aspartate aminotransferase (AST) > 5 X upper limit of normal (ULN) 4) pregnancy 5) breast feeding.

6) history of severe asthma.

### Intervention

Patients received IVM 200 Mcg single dose at the admission day as add on therapy to Iraqi Ministry of Health protocol for treatment of mild to moderate COVID-19 [HCQ 400mg BID for the first day then 200mg BID for 5 days plus AZT 500mg single dose in the first day then 250mg for 5 days]. We evaluated these patients for cure by clinical assessment and PCR swab testing. Nasopharyngeal or oropharyngeal swabs specimens were collected on days 1, 3, 5, 7, 9, 11, 13, 15, 17, 19, 21, and 23 for viral RNA detection and quantification till two successive days of negative PCR swab testing at least 24hrours apart. Virological testing was done at Alshifa’a Hospital Laboratory Center using ABI 7500Dx Real-Time PCR System instruments (Applied Biosystems), USA.

### Outcomes

The primary outcome was percentage of the cured patients within 23 days. Cure of the patients was defined by assessing proportion of patients who were symptoms free to be discharged from the hospital and included body temperature returned to normal for longer than 3 days, respiratory symptoms significantly improved, and 2 consecutive negative PCR test results from nasopharyngeal swabs at least 24 hours apart. The secondary outcomes were time to cure in both groups. Time to cure is evaluated by measuring time from admission of the patient to the hospital till discharge after being free of symptoms and negative PCR swab. Once nasopharyngeal and oropharyngeal swab viral PCR testing yielded negative results 2 times consecutively, no further testing was performed. Also safety outcomes included treatment-emergent adverse events, serious adverse events, and premature discontinuations of study were recorded if present.

### Sampling method and sample size calculation

A convenient consecutive sample of patients were enrolled in the study. The sample size calculated for this pilot trial was 30 patients : 15 in the active arm (IVM group) and 15 in the controls (SCA) according to pilot study sample size rule of thumb to get medium effect size of 0.3 ≤ δ/σ < 0.7 with a statistical power of 90%. (9)

### Statistical analysis

Statistical analysis was done using R packages software for IOS. The normality of continuous variables was analyzed using Shapiro Wilk test. Continuous variables were expressed as mean ± standard deviation (SD) if were normally distributed and median (interquartile range) if not normally distributed. Categorical variables were presented as number and percentages. Difference between normally distributed continuous variables was measured using Student’s t-test and Mann-Whitney U test if not normally distributed. Effect size for non normally distributed variables was measured using Vargha and Delany A test. Kaplan Meier survival curve analysis with and log rank testing was used. The standardized mean difference effect size is small if value 0.2 - 0.5; medium if value 0.5-0.8, and large effect size if value 0.8-1.4 P value less than 0.05 was considered statistically significant.

## Results

### Population characteristics

#### Participant flow

A total of 20 patients were screened for IVM add on group. Of those 4 patients were excluded: 2 of them due to severe COVID-19 and 2 had incomplete data and diagnosis. For comparison with the SCA, a total of 84 patients were screened for eligibility, of them 13 patients were excluded due to having severe COVID-19. The eligible patients in the controls were 71 patients. Two of them died during follow up and 69 completed their standard protocol of therapy according to the Iraqi ministry of health protocol of treatment of COVID-19 as shown in figure 1.

**Figure 1:**
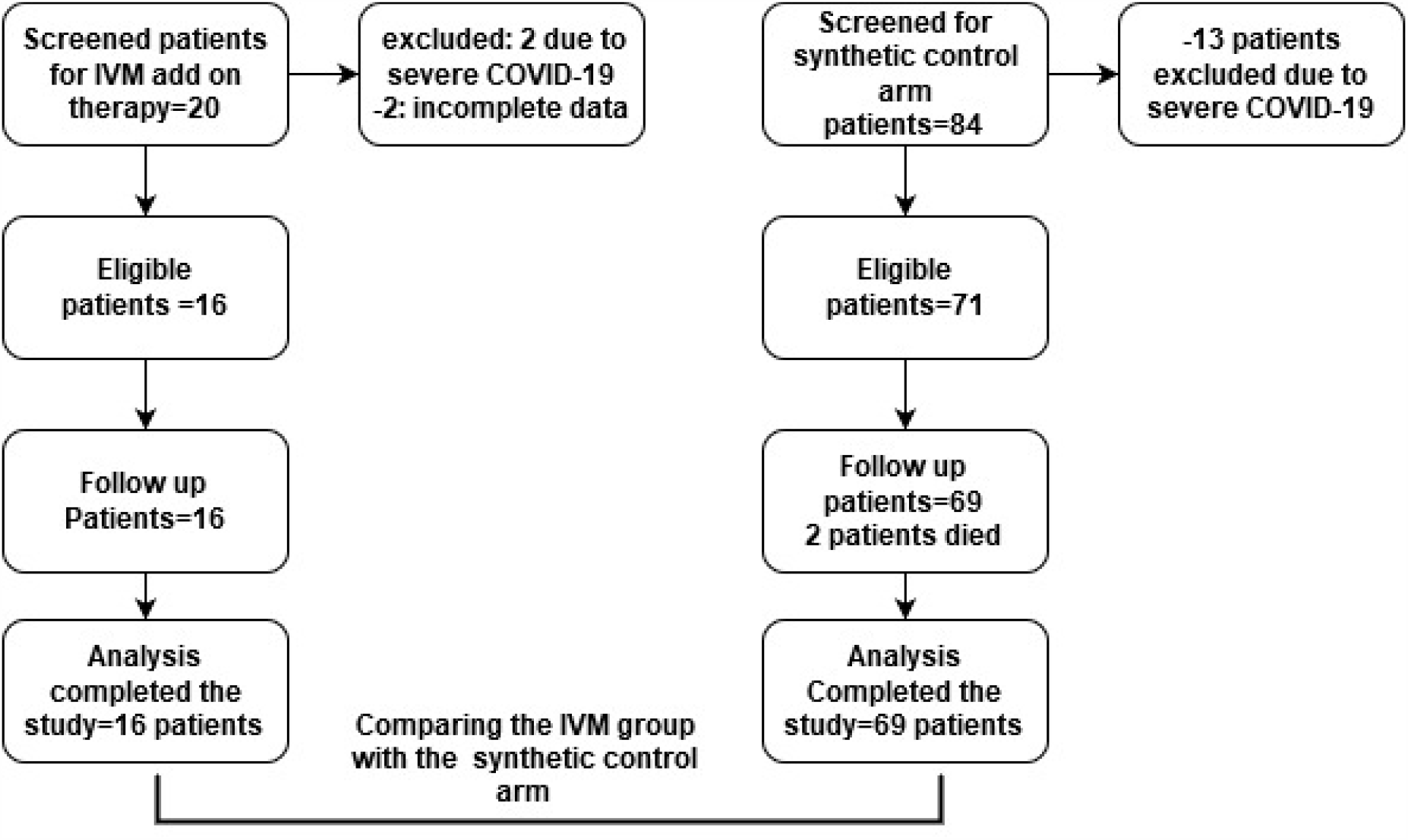
Flow chart of the study group.

#### Baseline characteristics

Table 1 shows that mean age ± SD of patients in the IVM group was 44.87 ± 10.64 years with a range of (28-60) years and for the controls was 45.23 ± 18.47 years with a range of (8-80) years. Majority of patients of IVM and controls were male [11(69%) versus 52 (73%), with male: female ratio 2.21 versus 2.7-1 respectively. Most cases of COVID-19 were mild in both groups [9(65%) in IVM versus 40(56%) In non IVM]. The most common clinical features in IVM group were cough 13(81 %), next fever 11(685), then shortness of breath 9(56%). Similarly, in non IVM group, the most common clinical features were: cough 55 (77 %), next fever 53 (74 %), then shortness of breath 44 (61%). Four patients in IVM group had underlying diseases: of them three had diabetes mellitus and hypertension and 1 had asthma, while in the non IVM, 29 (45%) had underlying diseases, of them 15 had diabetes mellitus, 14 had hypertension, and seven had asthma. There was no statistical significant difference between IVM group and the controls indicating no signiant sociodemographic and clinical confounders affected the study (p>0.05)

**Table 1:**
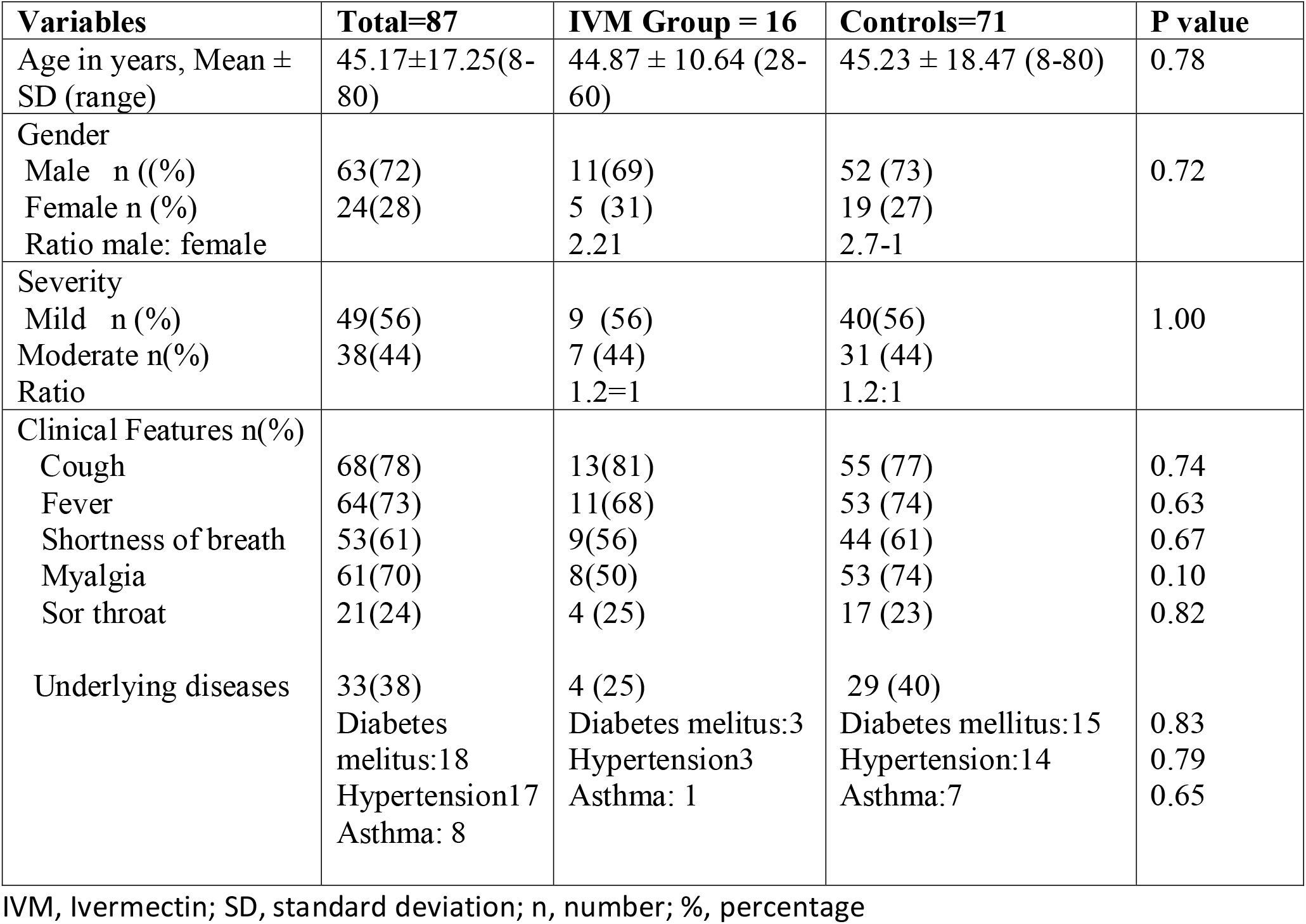
Baseline characteristics of IVM and Non IVM group

#### Outcomes and estimation

In Table 2, All the patients 16 (100 %) of IVM group were cured compared to 69 (97.2%) in the non IVM group. There were two patients died in the non IVM group.

**Table 2:**
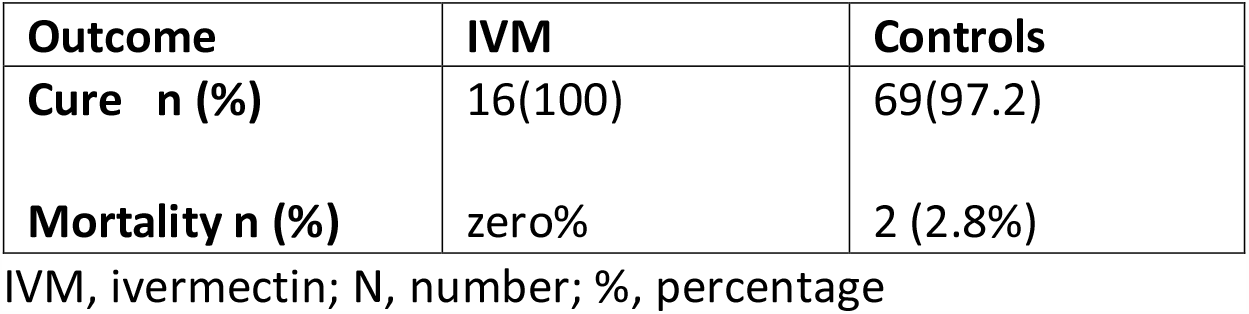
Percentage of cure rate IVM group and Non IVM group Cure rate

| **Outcome** | **IVM** | **Controls** |
| --- | --- | --- |
| **Cure n (%)** | 16(100) | 69(97.2) |
| **Mortality n (%)** | zero% | 2 (2.8%) |

IVM, ivermectin; N, number; %, percentage

The mean time to stay in the hospital was lower in IVM group compared with the controls and was statistically significant and clinically relevant (7.62 ± 2.75 versus 13.22 ±5.90 days, p=0.00005) with large effect size = 0.82) as in figure 2.

**Figure 2:**
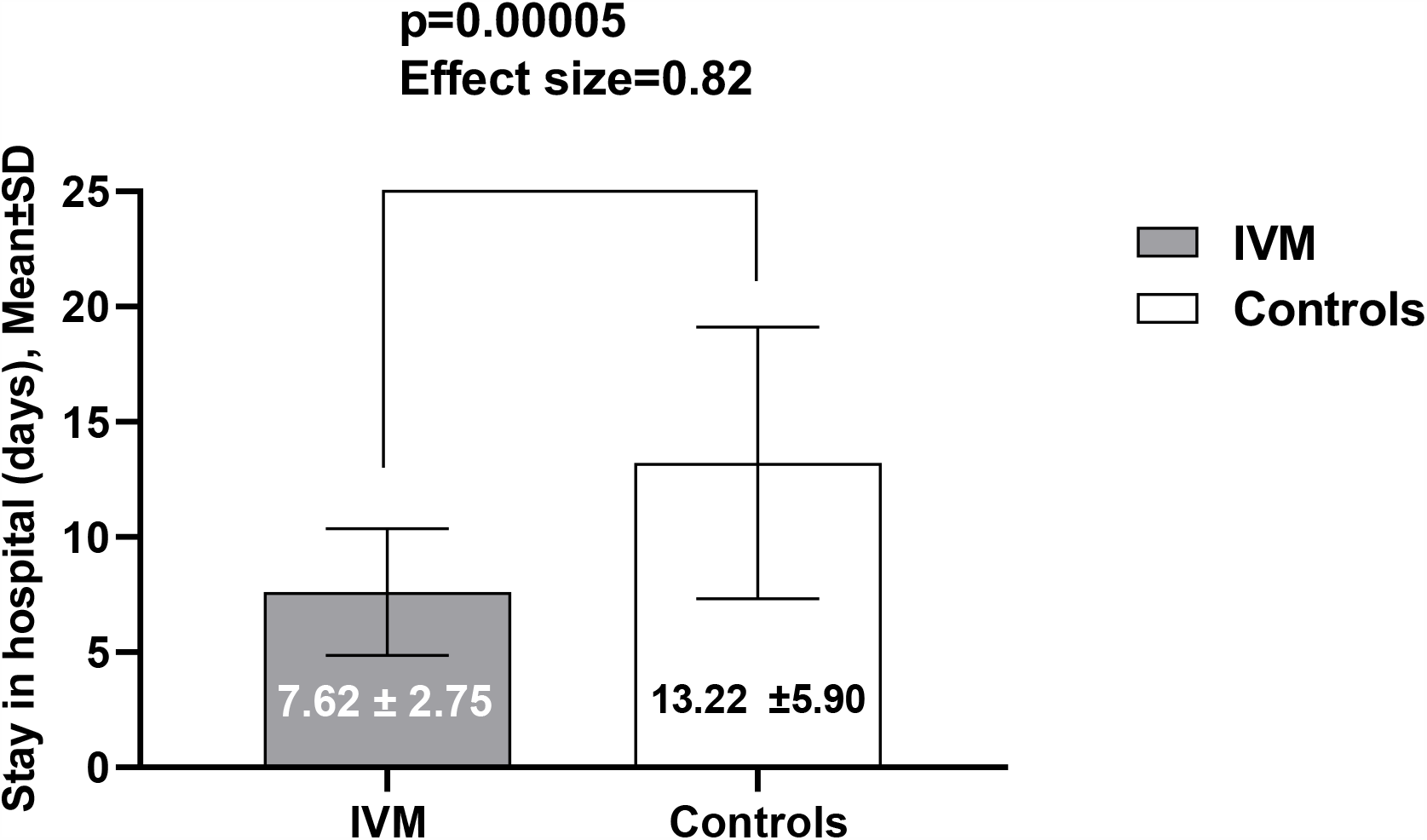
Mean stay days in hospital in IVM group compared with controls

The percentage of positive PCR patients with IVM group had significantly shorter time to become negative PCR compared to the controls. The median days of positive PCR in the IVM group was significantly lower than that of controls [7 (95% CI 6-11) vs 12 (95% CI 10-15), log rank test p<0.001 respectively] as in figure 3.

**Figure 3:**
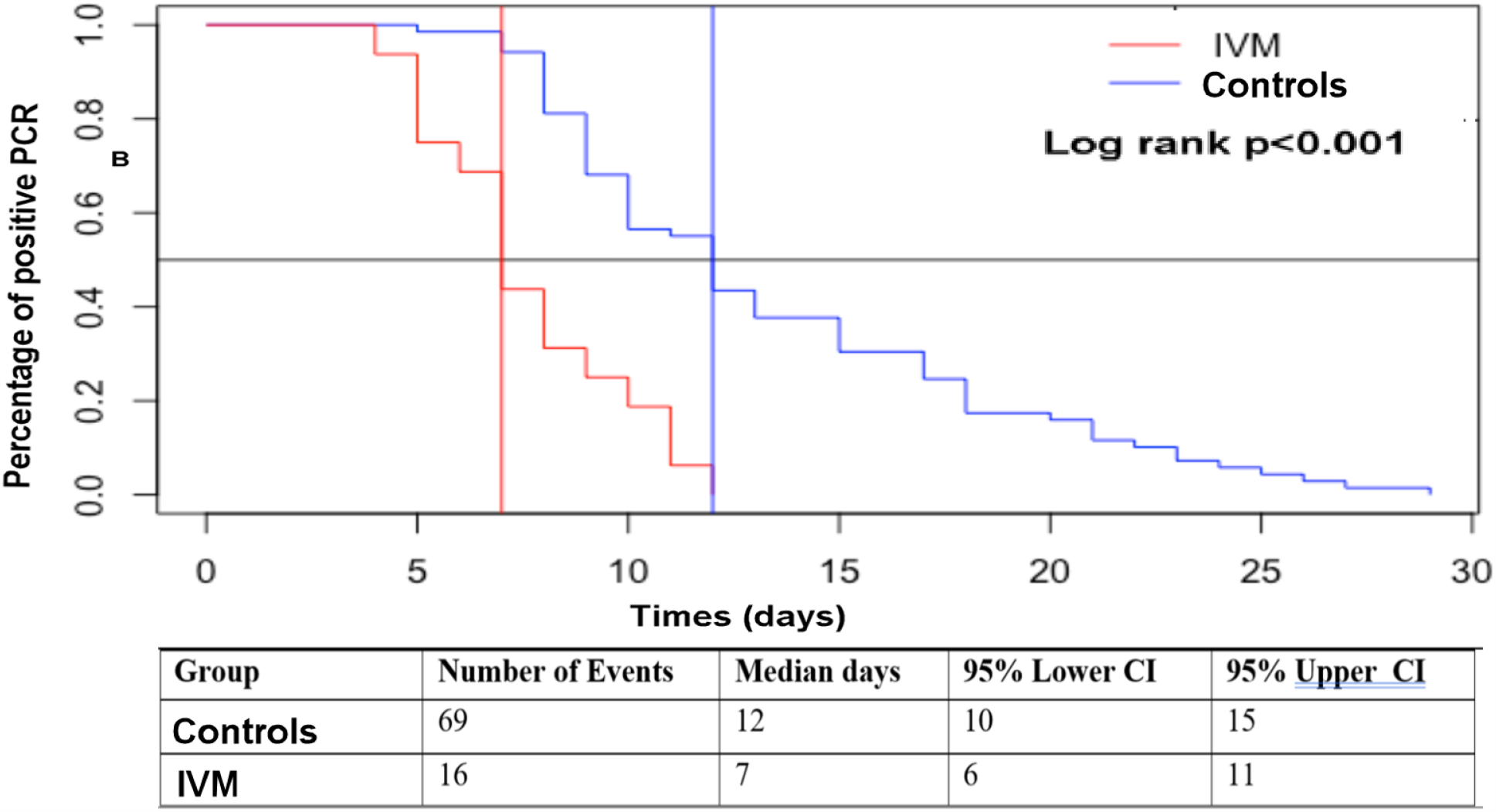
Kaplan Meier survival analysis curve for time to percentage of positive PCR in IVM group (n=16) versus controls (n= 69) (p<0.001, Log rank test). PCR, polymerase chain reaction; IVM, ivermectin; CI, confidence interval. The vertical red line represents the median days of IVM; The vertical blue line, represent the median days for controls.

No side effects have been found in the IVM group.

## Discussion

To date, no effective therapy has been shown for patients with COVID-19. This preliminary pilot study demonstrated for the first time that add-on use of IVM to HCQ and AZT had obvious higher cure rate, shorter hospital stay days compared with controls. In addition, there was no obvious reported adverse events.

Although data from several ongoing randomized controlled trials (RCT) for IVM will soon provide more informative evidence regarding safety and effectiveness IVM for COVID-19. The outcomes observed in this study with synthetic controls are the best available data regarding IVM use for COVID-19. In addition, during a pandemic with a potential death outcome, RCT become difficult to conduct and may be unethical. Randomization to placebo could carry a risk of serious consequences and even Food and Drug Administration (FDA) is encouraging the use of synthetic control for more innovative approaches (10).

According to the results of this study, all the patients 16 (100%) in the IVM group were cured compared to 69 (97.2%) in the controls. No similar study to compare with.

A Pilot observational study conducted by Gautret et al to assess the clinical and microbiological effect of a combination of HCQ and AZT in 80 COVID-19 patients with at least na six-day follow up reported that all patients recovered except three patients, one of them died and the other two admitted to the intensive care unit (11).

Another open label non randomized trial evaluated six patients taking HCQ and AZT compared with 14 patients on HCQ and 16 untreated patients from another center and cases refusing the protocol were included as negative controls reported 100 % cure rate for those on combined HCQ and AZT, 57.1 % on HCQ only, and 12.5% (p<0.001) (12)

Recently, in a large international, multicenter, observational propensity-score matched case-controlled study in 1,408 patients (704 received IVM in a dose of 150 mcg/kg and 704 that did not) showed that fewer patients of those requiring mechanical ventilation died in the IVM group (7.3% versus 21.3%) and overall death rates were lower with IVM (1.4% versus 8.5% with a corresponding HR 0.20, CI 95% 0.11-0.37, p<0.0001).They concluded an association of IVM use with lower in-hospital mortality and suggested a potential survival benefit of IVM in COVID 19 (13)

In contrast, Molina et al reported in a letter to editor that no evidence of rapid antiviral clearance or clinical benefit with the combination of HCQ and AZT in patients with severe COVID-19 infection despite a reported antiviral activity of HCQ against COVID-19 in vitro and suggested ongoing randomized clinical trials with HCQ to provide a definitive answer regarding the alleged effectiveness and its safety (14). The variation in the results in those studies may be related to their study design, included severe patients, small sample sizes, inaccurate sampling method, and short follow up with repeated qualitative PCR assay.

In a randomized, controlled trial of lopinavir-ritonavir in adults hospitalized patients with severe COVID-19 reported no significant benefit was observed with lopinavir-ritonavir treatment beyond the standard care. However, in the modified intention to-treat analysis, which excluded three patients with early death, the between-group difference in the median time to clinical improvement (median,15 days vs. 16 days) was significant, albeit modest. The overall cure rate was 77.9% in that trial. The explanation of high mortality rate possibly was related to the severely ill population enrolled in that study (15)

Moreover, in another recent trial, a preliminary report of compassionate use of remdesivir for small cohort of patients with severe COVID-19 showed an observed clinical improvement in 36 of 53 patients (68%) and overall mortality was 13% over a median follow-up of 18 days. The relatively high mortality rate and less cure in their results may be related to the sever type of COVID-19 in their patients. (16).

Another noteworthy observation in this study was the time to stay in the hospital. The results revealed that the mean time to stay in the hospital in IVM group was 7.62 ± 2.75 days compared with 13.22 ±5.90 days for the controls which was statistically significant (p=0.00005) and clinically relevant with large effect size (Effect size=0.82). This mean shorter time to recovery and early time to discharge patient to home in those taking IVM which will help to provide more beds for another patients who need it and this is practically important in this pandemic disease. Up to our knowledge there is no other study to compare with it.

The time of rapid and full viral clearance was controversial in literatures regarding combination of HCQ and AZT. One study reported that a 100% viral clearance in nasopharyngeal swabs in 6 patients after 5 and 6 days of the combination of HCQ and AZT (17). Another study observed that 10 patients (not done on patient who died) out of 11patients were still positive at day 6 after initiation of treatment on repeated nasopharyngeal swab using a qualitative PCR assay (nucleic acid extraction using Nuclisens Easy Mag®, Biomerieux and amplification with RealStar SARS CoV-2®, Altona) (13).

In addition, a new study from China in individuals with COVID-19 found no difference in duration of hospitalization at 7 days with or without HCQ (18). The difference in the study design and short follow up with repeated PCR may related to the variation in the day of cure in those studies.

On comparing the mean cure days of patients in our study to the mean recovery of other medications, we found two recent studies. The first study reported the median duration of recovery with 95% CI of patients receiving remdesivir compared with PBO was of variable ranges from 11 (9–12) days in remdesivir versus 15 (13-19) days in PBO to unestimated days in remdesivir versus 28 days in PBO (16). This variation can be explained according to the severity score of the disease on the ordinal scale of intention to treat analysis they used in their study, those patients with score four or less had less duration of cure compared to those with score seven which was an expected finding. The second study compared lopinavir/ritonavir to the standard care and showed that the median time to clinical improvement in days (interquartile range) was 16.0 (13.0 to 17.0) days in lopinavir/ritonavir group versus 16.0 (15.0 to 18.0) in the standard care group which was not significant (15).

In the current short-term study, no new safety signals were detected apart from patients’ manifestations on hospital admission. There were no obvious immediate or late adverse events that occurred during treatment, and no serious adverse events that necessitate discontinuation of treatment However it is challenging to attribute any new abnormal complications in the patients whether due to IVM use or the disease itself.

The age, gender, severity of the disease, and the comorbid diseases in the IVM group were not statistically significantly different from the controls. This indicate that these variables were not significant confounders that affect our results and the findings we got were mostly due to the effect of IVM rather than those confounders.

The mechanism of better response in IVM group compared with the controls in this study may be due to the possible synergistic action of these three drugs (IVM, HCQ, and AZT). It was reported that HCQ and IVM were known to act by creating the acidic environment and inhibiting the importin (IMPα/β1) mediated viral import. AZT was found to act similar to the HCQ as an acidotropic lipophilic weak base. All the three categories of drugs seemed to potentially act against COVID 19. infection (19).

This study had some strengths. It is the first externally controlled pilot trial; performed in a public hospital, strict exclusion and inclusion criteria, and presented for the first time assessment of the effectiveness and safety of add on use of IVM to HCQ and AZT. However, this study has some limitations, including its small sample size; single center design, short time for the study, and being nonrandomized.

In conclusion, this study showed that adding IVM to HCQ and AZT had a better cure rate and shorter time to stay in the hospital compared with controls. In addition, it was relatively safe without observable safety signals. These findings may suggest using IVM as an add on therapy to protocols used for treatment of COVID-19. However, these results are needed to be validated in a larger prospective follow up study. The results of the study must not be considered conclusive since unknown confounders cannot always be reliably accounted and we recommend further studies. A national multicenter randomized study is planned to perform in different provinces of Iraq using IVM alone since HCQ is temporary withdrawn from the COVID 19 therapy according to the WHO advice.

## Data Availability

Data will be available when required

## Acknowledgements

We thank all the participant patients in the study. Also, we thank the medical staff who helped us in data collection.

